# Community mobilisation approaches to preventing and reducing adolescent multiple risk behaviour: a realist review protocol

**DOI:** 10.1101/2021.03.16.21253705

**Authors:** Laura Tinner, Deborah Caldwell, Rona Campbell

## Abstract

**Background:** Adolescent multiple risk behaviour (MRB) continues to be a global health issue, contributing to the burden of non-communicable diseases. Most interventions have focused on the proximal causes of adolescent MRB such as peer or family influence, rather than targeting the wider environmental or structural context. There is increasing recognition that community mobilisation approaches that extend beyond individually-focused educational programmes could be beneficial for adolescent health. Despite this, there are gaps in the current literature, theory and implementation that would benefit from a realist approach due to the suitability of this methodology to analysing complex interventions. In this protocol, we outline our study that aims to understand ‘how, why, for whom and in what circumstances and time periods do community mobilisation interventions work to prevent and/or reduce adolescent multiple risk behaviour?’

**Methods:** A realist review was chosen as the most suitable review method as it is theory-driven and seeks to understand how, why and for whom interventions work to produce intended and unintended outcomes. A six-stage iterative process is outlined, which includes initial development of a programme theory, systematic searching, study selection and appraisal, data extraction and data synthesis. We will engage with stakeholders at different stages in this process to aid the development of the programme theory.

**Discussion:** The goal of this realist review is to identify and refine a programme theory for community mobilisation approaches to the prevention and/or reduction of adolescent multiple risk behaviour. Our aim is that the findings surrounding the programme theory refinement can be used to develop and implement adolescent multiple risk behaviour interventions and maintain collaboration between local policy makers, researchers and community members.

**Registration:** This realist review is registered on the PROSPERO database (registration number: CRD42020205342).

## Background

### Adolescent health risk behaviours

Health risk behaviours such as tobacco smoking, hazardous alcohol consumption, antisocial behaviour, physical inactivity and unprotected sexual intercourse are global health issues, which are commonly initiated and become habitual in adolescence (1, 2). Adolescents who engage in one risk behaviour are likely to engage in others (3, 4), leading to increased public health interest in multiple risk behaviour (MRB), which refers to the occurrence of two or more risk behaviours directly or indirectly related to health (5, 6). MRB has been found to be associated with a number of adverse health and social outcomes such as poor educational attainment (7), obesity, depression and anxiety in adulthood (8), cancers and premature mortality (7, 9). This has, in turn, led to public health interventions that address multiple as opposed to single behaviours (10).

Most interventions addressing adolescent MRB have focused on the proximal causes such as peer or family influence, rather than targeting the wider environmental, social or structural context (11). For instance, two Cochrane systematic reviews have assessed the impact of individual, family and school-level interventions on adolescent multiple risk behaviour (10, 12). One of those reviews found mixed evidence, concluding that school-based universal interventions are potentially effective in ‘preventing engagement in tobacco use, alcohol use, illicit drug use, and antisocial behaviour, and in improving physical activity among young people, but not in preventing other risk behaviours’ (12). The authors highlighted that there was no strong evidence of benefit for family-level or individual-level interventions across the risk behaviour outcomes investigated (12). The interventions included in this review were predominantly educational programmes. The effectiveness and equity of these “downstream” interventions has been questioned (13) because health risk behaviours rarely have a single cause and occur in complex socio-cultural contexts (14). As such, there is increasing recognition that structural changes that extend beyond individually-focused educational programmes could be beneficial for adolescent health (14, 15).

### Community mobilisation interventions

Recognition that decisions about health risk behaviours are made within a broad social context has led to the development and implementation of community-engagement interventions (16). There is an extensive range of types of community-engagement public health interventions, varying in the extent to which they emphasise community involvement in determining and delivering the programmes (16). ‘Community mobilisation’ interventions are one such type that work to engage community members to ‘take action towards achieving a common goal’ (17) and have gained traction as a strategy for addressing complex and multifaceted problems (18). Community mobilisation is a collaborative public health effort that is defined by the inclusion of a community coalition made up of diverse stakeholders (such as schools, businesses, residents, youth groups, emergency services and religious leaders) (11). These stakeholders critically analyse the root causes of local problems, identify an array of potential solutions, develop multi-sector partnerships, and implement multi-component strategies for creating local change and more health-promoting environments (14).

Community mobilisation efforts explicitly seek to affect community-level influences through changes of policies, practices, organisations and other features of the social or physical environment that may impact on the health outcome or behaviour (19), signifying a shift away from individual behaviour change to a focus on the social determinants of health (20). However, these approaches may still include components which address individual behaviours (e.g. health promotion programmes within schools), but they seek to combine these with other structural factors as part of a package of measures that are chosen and monitored from community stakeholders.

There is systematic review evidence suggesting that higher levels of community involvement within an public health intervention is linked to more beneficial effects and positive trends across a range of outcomes (21). There is also some evidence to support the role of community mobilisation efforts in preventing health risk behaviours. For instance, such interventions have resulted in reductions in high risk alcohol consumption and alcohol-related injuries (22); alcohol impaired driving (23); uptake of smoking in young people (16) and youth violence (24). Researchers have highlighted that with adequate resources and training, support from within the community and adoption of evidence-based strategies, community mobilisation approaches have promise as an effective vehicle for addressing adolescent multiple risk behaviour (19). Further, community-mobilisation efforts are also thought to be well suited to achieving health equity (25), due to ‘shared decision making’ (26) and the incorporation of ‘upstream’ or structural elements (27), but this has yet to be explored in relation to adolescent multiple risk behaviour interventions.

There are also significant challenges in implementing and evaluating such approaches, which is unsurprising given the dynamic set of social interactions and relational complexity one might expect in community-centred interventions (28). These implementation challenges include lack of community interest and long-term engagement, design inadequacies, inflated expectations, and weakness in planning and implementation of the interventions (19, 29, 30). Tensions and different expectations between scientists and community members as well as the practical difficulty in managing multiple components and stakeholder interests have also been cited as issues (11).

Evaluation is equally challenging (31), which is reflected in the lack of empirical evaluations of structural interventions such as community mobilisation compared to those focused at the individual level (14). There is uncertainty around how long it might take to see an impact on behaviours, although it is expected to be a lengthy process. Even if effects are identified, the chain linking any changes in health risk behaviours to the mobilisation efforts is so long and complex that causal attributions become complicated (14). The challenges in evaluating and implementing community mobilisation interventions has meant they are often evaluated through methods such as quasi-experimental studies in addition to randomised controlled trials (RCTs), meaning that they have been missing from systematic reviews such as the aforementioned adolescent MRB review (12). Consequently, the evidence base for community mobilisation efforts is mixed in terms of producing desirable outcomes on a community and individual level.

We aim to address the gap in the literature through investigating community mobilisation interventions aimed at preventing and/or reducing adolescent multiple risk behaviour. There is a strong rationale for an alternative review approach that speaks to the complexities and challenges surrounding the delivery and evaluation of community mobilisation efforts. Further, we are concerned with moving beyond assessing effectiveness of public health interventions to synthesise existing knowledge and articulate *how* community mobilisation interventions work for adolescent health. Therefore, a realist review was chosen as the most appropriate methodological approach. Realist reviews are theory-driven approaches to evidence synthesis, incorporating diverse data sources to provide insight into the underlying mechanisms and contexts in which the interventions work (32). Realist reviews are ideal for examining social interventions, particularly those in community settings as it is recognised that programmes are rarely delivered in the same way or have exactly the same outcomes, due to contextual factors that can never be fully controlled (33).

Realist inquiry is thus ‘increasingly recognised as an effective process for consolidating evidence and learning from complex social processes and interventions’ (34), with successes in public health and community development (35, 36). Therefore, our realist review aims to contribute to the current adolescent multiple risk behaviour evidence base, which has largely focused on effectiveness of interventions through traditional systematic reviews. Further support for our intended approach comes from a recent PhD thesis which used an adapted realist approach to assessing adolescent multiple risk behaviour programmes, combining realist evaluation with primary data collection (37) (38). The authors did not specifically focus on community mobilisation as an intervention and included fewer health risk behaviours than we do here. This protocol describes our realist methodological approach and intended procedures in the sections that follow.

This realist systematic review is registered on the PROSPERO database (registration number: CRD42020205342). A PRISMA-P check list is included as a supplementary file.

## Methods

### Review aim

Our aim is to use a theory-driven evidence synthesis to assess how and why community mobilisation interventions work/do not work to prevent or reduce adolescent multiple risk behaviour and in what contexts. We are additionally interested in the question of ‘who’ these interventions work for, in order to understand the impact of these types of interventions upon existing health inequalities through investigating whether the interventions are beneficial to disadvantaged communities. Although the focus of the review is adolescent multiple risk behaviour, we may draw on wider literature to understand the goals of community mobilisation approaches and the mechanisms by which it is hoped these are achieved. An additional objective of our review is to develop transferable learning about community mobilisation approaches in public health research and adolescent health interventions. The realist review will be guided by the following sub-questions:

1. What are the outcomes of community mobilisation interventions targeting adolescent multiple risk behaviour?
2. What are the mechanisms, acting at the individual, community and societal levels through which community mobilisation interventions produce outcomes?
3. What are the key contextual influences that determine whether the mechanisms produce both intended and unintended outcomes?

## Realist review methodology

Realist review methodology is a theory-driven, interpretive approach to evidence synthesis (38) developed by the work of Pawson et al (39-41). It has gained increasing popularity in addressing the challenge of ‘what works, for whom, under what circumstances and in what time period’ and is considered especially salient when data are complex, multi-layered and there is a need to understand complex relationships, interdependence, and mechanisms (42). A core component of realist reviews is to develop ‘middle-range realist programme theory’ (43) that explains how an intervention ‘works’ within what contexts. Realist reviews allow for exploration of complex topics and the inclusion of a wide body of quantitative, qualitative and mixed methods evidence to develop and refine theory (38). Therefore, it is suited to multi-component community-based interventions, for which evaluations may include a range of different data and be published in grey literature.

Realist reviewers view ‘causation’ as generative, which means that the manifested world is generated (i.e. caused) via underpinning mechanisms (32). They identify where an intervention, under certain contextual conditions (C), triggers a mechanism (M) to achieve a given outcome (O) (38). This CMO configuration is central to analysis and theory development, viewing mechanisms as the integral link between contexts and outcomes. The aim of realist review methodology to move beyond measuring effectiveness of interventions, toward explanation of how and why an intervention works is a key strength that separates it from traditional systematic reviews. Dalkin et al (44) conceptualise a mechanism as a resource or reasoning, which triggers an outcome, but may only be active in certain contexts. Under certain contextual conditions, the mechanisms are triggered, while in others they ‘fire’ to a lesser degree or not at all (44). Realist review methodology is highly applicable to complex public health interventions and is an approach that can build ‘common ground’ between researchers and policy makers through providing accessible recommendations on how interventions might be delivered in different contexts (41).

We have conceptualised multi-component community mobilisation approaches as complex interventions in which outcomes and mechanisms will be context sensitive (39). Therefore, the realist review approach will allow us to investigate in what contexts community mobilisation interventions are effective. This realist review will follow the practice guidelines outlined by the Realist and Meta-narrative Evidence Synthesis: Evolving Standards (RAMESES) framework (40, 41).

## Study design

This review is structured around the five review stages outlined by Pawson et al (39) and has been informed by other realist review protocols in the field (38, 45). Figure 1 is a diagram of the review process adapted from Power et al (38).

**Figure 1:** Summary of stages of realist review adapted from Power et al (38). This depicts the steps for developing the initial programme theory, searching for evidence and synthesising the data with the input of key stakeholders. Retroduction refers to inferences made through interpreting the data about the underlying causal mechanisms.

## Stages of realist review

### (1) Locating existing theories

The first step in a realist review is to conduct scoping searches that begin to identify theories that might explain how community mobilisation interventions may work to address adolescent multiple risk behaviour. The search will include academic databases (MEDLINE, PubMed, Web of Science), UK health websites and grey literature databases (OpenGrey, the King’s Fund, The Health Foundation) as well as Google Scholar. Broad search terms will be used at this initial stage (e.g. “community mobilisation”, “community coalition”, “youth”, “adolescence”, “health risk behaviour”, “substance use”, “antisocial behaviour”) and back and forth citation tracking will be utilised until we develop a core set of empirical studies to help build the initial programme theory framework (40). This initial search is not designed to be exhaustive: this stage in the theory development is expected to be a ‘rough starting point’ that will be refined throughout the realist review process (43).

It is advisable to include the expertise of those delivering or evaluating the interventions. At this stage we will engage with key stakeholders identified through the literature to provide guidance on the development of the programme theory. For example, they may provide insight into the different contexts and mechanisms that impact on adolescent risk behaviour outcomes from their experience in the field. These stakeholders may also highlight other relevant studies or individuals we should engage with to further develop the programme theory. When we have developed an initial programme theory, we will move onto stage 2 and the more structured and systematic searching.

### (2) Search strategy

In Stage 2 we will conduct more formal searches, which will be informed by the initial programme theory development in stage 1. The objective in this stage will be to identify literature and evidence capable of informing the refinement of a more detailed programme theory (45). We will develop search terms from the initial background search in Stage 1 and discussions with a subject librarian, leading to systematic searches being undertaken to collect evidence to refine the programme theory. We will include the following databases:

PubMed; MEDLINE; PsycINFO; Web of Science; CINAHL; Sociological Abstracts. Grey literature will also be searched on OpenGrey and on external expert organisations and charity websites. ProQuest will be searched for unpublished theses and dissertations. Google Scholar will be used for citation searching as well as reference lists of relevant papers.

Search terminology and syntax will be informed by the initial programme theory identification, known literature and collaboration between the research team and a subject librarian. We will draw on the search terms used in previous systematic reviews on individual-level adolescent multiple risk behaviour interventions. Search terms will include MeSH terms and free text related to “community mobilisation”, “adolescence” and a range of multiple health risk behaviours. No date restrictions will be used and only studies in the English Language will be assessed for eligibility.

While formalised and systematic, the sampling approach in realist reviews remains purposive to answer specific questions and develop theories (39). Therefore, the process will likely be iterative and need to be repeated (39), with back and forth citation tracking remaining a key part of the iterative search strategy (46). Corresponding authors of selected articles may also be contacted for further examples that may be relevant to the question. The search terms and strategies will be documented in a log-book as the review progresses.

### (3) Study selection

We will use the following inclusion criteria to determine if a document is likely to contribute to the programme theory development:

- Type of intervention: Community mobilisation must form a core part of the intervention, most commonly identified by the development of a community coalition group involving a diverse range of community stakeholders as identified above. In many cases, the community coalition will select intervention components from a ‘menu’ of strategies and adapt them to fit local needs. As mentioned, although these types of interventions derive from the desire to move away from individual behaviour change, they are also likely to include a range of “upstream” and “downstream” components, which may include education delivery. The key criterion remains that the intervention should include community mobilisation at its core, and should include at least two components (e.g. an educational programme and local policy enforcement).
- A range of document types, study designs and data types may be relevant to the development of the initial programme theory. All intervention evaluation study designs and data types, from all time periods, may be included in the review to test and refine the programme theory (e.g. randomised controlled trials (RCTs), quasi-experimental studies, case studies).
- Participants: the community mobilisation efforts should be targeted (at least predominately) at young people age 10-19 years, although this age range remains flexible^1^. Adult stakeholders (such as parents, community members, school staff) will likely be included in the community coalition but should not be the focus of the intervention. All sampling decisions will be transparently reported.
- Aim of intervention: the intervention should have a primary focus on prevention and reduction of adolescent multiple risk behaviour and include at least two health risk behaviours from a wide range including: regular tobacco smoking; regular alcohol drinking; binge drinking (alcohol); cannabis use; recent or regular illicit drug use; risky sexual behaviours; anti-social behaviour and offending; vehicle-related risk behaviours (e.g. cycling without a helmet; not using a car seatbelt, joy riding); self-harm; gambling; unhealthy diet; and physical inactivity.
- Outcome measures: Primary outcomes of interest include reduction and/or prevention of the wide range of multiple risk behaviours mentioned above. A range of additional medium- and long-term outcomes within health and social domains are expected given the number of health risk behaviours the interventions can cover as well as the multi-component nature of these types of interventions. Secondary outcomes of interest in this review include: Education and employment: educational qualifications; truancy and school exclusion; employment; not being in education, employment or training (NEET); Crime: Criminal record/offending; long-term addictive behaviours; Gambling; Teenage pregnancy or parenthood; Sexually transmitted infections; Injuries; Morbidity (e.g. Hepatitis C, HIV, anxiety and depression, obesity, type II diabetes, fatty liver disease, liver cirrhosis); Suicide/self-harm; and Premature mortality. Realist reviews are interested in intended and unintended outcomes related to the context, mechanism, and outcomes of the intervention therefore other unknown outcomes may become relevant to the programme theory development.

Studies will be excluded if they relate to any of the following:

- A single component intervention (such as an educational programme) that is delivered in the community but does not incorporate community mobilisation as we have defined it.
- Interventions targeted at participants outside of the age range. Some interventions may include other populations, but youth should be the primary focus.
- Interventions aimed at preventing and reducing a single adolescent health risk behaviour (e.g. alcohol misuse).
- Clinical and pharmaceutical interventions including ‘community outreach’ services such as the provision of mobile clinics.
- Studies not described in the English language.

We will use Raayan (QCRI) software for screening and management of the studies at this stage in the review. The RAMESES guidelines will be used to appraise the studies (41). The selection of evidence will be made based on judgements around their *relevance* (contribution to the programme theory development and refinement) and *rigour* (credibility and trustworthiness of methods) (45, 47). Any exclusions based on these appraisals will be documented.

### (4) Data extraction

Study characteristics will be extracted into a table to provide a descriptive overview of the types of community mobilisation interventions included, based on a ‘bespoke’ set of data extraction forms informed by the relevant literature (39). Realist reviews are structured through Context-Mechanisms-Outcome (CMO), comparable to PICO for traditional systematic reviews (42). Context, mechanisms and outcomes are extracted during the realist review and can be conceptualised as the ‘data’ that support evidence to support, reject or refine the programme theory (33). However, this process is not as rigid as with traditional systematic reviews and different sources may provide different information that contributes to the programme theory development. We will extract the following information:

- Study details: authors, year of publication, country of intervention delivery, study aims, study design, participant characteristics, quality appraisal.
- Context: background of the intervention, aims of the intervention, type of intervention, setting (e.g. type and size of community), age range, number of components, policy context for the area, historical context.
- Mechanism: descriptions of the processes through which the intervention influenced outcomes, who the intervention worked for and who it did not, author-identified mechanisms.
- Outcomes: adolescent multiple health risk behaviour outcomes but also a range of health and social outcomes mentioned previously.
- Additional study information and researcher comments.

The above is not an exhaustive list and information on the mechanisms and context will be expected to change through experience with the studies and input from expert stakeholders. Any disagreements on extracted data will be resolved through discussion with the research team. Realist reviews assimilate information more through note taking and documents are scoured for ideas about how the intervention might work (39). To approach the more complex and iterative process of examining study sources, we will follow the guidance from Pawson et al (39) and will also upload the documents to NVivo for organising and coding to aid development of the programme theory and to keep a record of our procedures (45).

### (5) Data synthesis

The goal of data synthesis in a realist review is to consolidate the data from the previous steps to refine the initial programme theory (45). Analysis will involve interpretation of the researchers and judgement of the data. Coding the data will involve deductive (informed by the initial programme theory), inductive (emerging from the data within the identified interventions) and retroductive approaches (inferences made through interpreting the data about the underlying causal mechanisms) (45). These approaches will guide the review to interpret and explain the findings and outline the contextual conditions and mechanisms that may need to be present for outcomes to occur. Data to inform our interpretation of the relationships between the contexts, mechanisms and outcomes will be analysed within and across the documents. For example, mechanisms inferred from one document will be applied to other documents to test if they can explain the way contexts influence outcomes in another intervention (45). This interpretive process will lead to programme theory development of community mobilisation approaches to preventing and reducing adolescent multiple risk behaviour.

### (5) Refine programme theories

The final product of a realist theory is not a statement of effectiveness, but a refinement of middle-range theory that answers the questions of what works, for whom, under what circumstances, in what time period, why and how? (48). Evidence may include primary outcome data, but also rich description that conveys the contextual elements and interpretation of the interactions between the context, mechanisms and outcomes by the researchers (48). Stakeholders involved in stage one will be contacted again, with the potential for the inclusion of newly identified individuals, for input into the final programme theory(ies). The aim of this process is to confirm that the programme theory makes sense to those involved, in order to enhance our ability to make practice recommendations from our findings (39). If needed, we will re-scrutinise elements of the review based on the stakeholder involvement. We intend to develop a final logic model that will visually present the programme theory/theories and the relationships between the CMO.

## Discussion

The realist review approach will allow us to explore the relationships between contexts, mechanisms and outcomes and synthesise evidence surrounding adolescent multiple risk behaviour interventions that incorporate community mobilisation. We seek to gain a greater understanding of ‘what works, for whom, in what circumstances, in what time period and why’, in terms of preventive interventions to improve adolescent health and reduce inequalities. Our aim is that the findings surrounding the programme theory refinement can be used to develop and implement adolescent multiple risk behaviour interventions and maintain collaboration between local policy makers, researchers and community members. We will recognise any limitations to our study and the realist review approach in the final synthesis. The dissemination of the findings of this review will follow the RAMESES reporting guidelines (40, 41).

## Data Availability

This is a protocol so no available data yet

## Abbreviations

MRB: Multiple risk behaviour
NEET: not being in education, employment or training
HIV: human immunodeficiency virus
RCTs: Randomised Controlled Trials
CMO: Context, Mechanism, Outcome
RAMESES: Realist and meta-narrative evidence syntheses: evolving standards

## Declarations

### Ethics approval and consent to participate

Not applicable.

### Consent for publication

Not applicable.

### Availability of data and materials

Not application.

### Competing interests

The authors declare no competing interests.

## Funding

This work is funded through LT’s post-doctoral post, which is funded by Bristol, North Somerset and South Gloucestershire Clinical Commission Group (CCG) Research Capacity Funding via RC’s Senior Investigator Award. The funder had no involvement with the development of the protocol.

## Authors’ contributions

LT, DC and RC developed the project idea, chose the realist review approach and developed the inclusion criteria. LT wrote the first draft of the protocol. All authors contributed to writing and refining the protocol and will be involved in the review. LT is the corresponding author.

## Acknowledgements

Not applicable.

## Footnotes

1 For instance, a ‘pre-teen’ intervention targeting children age 9-12 would be included as the population are only one year out of our pre-specified age range of 10-19 years. However, an older adolescent/young adult intervention targeting individuals aged 18-25 would not be included as only two years (18 and 19) of the eight year range addressed by the intervention falls within our age banding.

## Notes

### Competing Interest Statement

The authors have declared no competing interest.

### Clinical Trial

It is a realist review protocol, for which the PROSPERO ID is: CRD42020205342

### Funding Statement

This work is funded through the lead author post-doctoral post which is funded by Bristol, North Somerset and South Gloucestershire Clinical Commission Group (CCG) Research Capacity Funding via a Senior Investigator Award. The funder had no involvement with the development of the protocol.

### Author Declarations

University of Bristol

